# Enhancing the Reliability of Resting ECGs via Deep Learning–Driven Motion Artifact Detection

**DOI:** 10.1101/2025.08.26.25334522

**Authors:** S.A. Nanayakkara, R.G.N. Meegama

## Abstract

This study presents a novel two-stage framework to enhance the reliability of resting electrocardiogram (ECG) signals by addressing motion artifacts that often compromise diagnostic accuracy. In the first stage, motion artifacts are mitigated using stationary wavelet transform coupled with Savitzky-Golay filtering, effectively preserving critical ECG morphological features such as the QRS complex. The second stage employs a deep convolutional neural network to classify ECG signals as either usable or artifact-corrupted, achieving a classification accuracy of 98.76%. Utilizing a 12-lead ECG dataset from PhysioNet, the proposed unified CNN model outperforms individual lead-specific models, offering superior computational efficiency (1.6 seconds vs. 21.7 seconds for predictions) and reduced storage requirements (1 GB vs. 15 GB). The approach demonstrates high sensitivity (98.74%) and specificity (98.77%), ensuring robust detection of noisy signals. By integrating advanced preprocessing with deep learning, this framework enhances ECG signal clarity, reducing the risk of misdiagnosis in clinical settings.

## 1. Introduction

The ECG is a vital tool for diagnosing various heart abnormalities [1]. By placing electrodes on specific locations on the skin, ECG measures the electrical activity associated with the heart muscle, which is propagated through various tissues to the skin’s surface. The resulting ECG waveform, consisting of P-wave, QRS-complex and T-wave, provides valuable information about the heart’s function, enabling medical professionals to detect potential issues [2].

The cardiac conduction system, which includes the sinoatrial (SA) node and atrioventricular (AV) node, plays a crucial role in maintaining the heart’s rhythm and the proper function 3, 4. However, despite the potential of ECG to identify heart diseases, a clear and uninterrupted signal is necessary for accurate diagnosis. Motion artifacts, which result from involuntary muscle movements and skin stretching, can distort ECG signals and complicate interpretation [5, 6].

Researchers have long sought effective methods to reduce motion artifacts and extract clean ECG signals. Traditional mathematical models and standalone machine learning approaches have struggled to achieve satisfactory results due to the low Signal-to-Noise Ratio (SNR) in ECGs. Moreover, interpreting ECG data requires expertise, further limiting the applicability of these methods.

There are various types of ECGs, categorized based on the method of acquisition and the purpose of the test. These include resting ECG, exercise stress test (EST), ambulatory ECG (Holter monitoring) and event monitoring. Resting ECG, as the name implies, is performed while the patient is at rest, typically lying down. This type of ECG is commonly used as the first step in diagnosing heart diseases and assessing the heart’s overall function.

Resting ECG is selected for this study because of its widespread clinical use and vulnerability to motion artifacts, even when patients are at rest. As noted earlier, involuntary muscle activity and skin stretching can introduce noise into the ECG signal during rest. These artifacts may cause misdiagnosis or necessitate further testing, leading to inconvenience for both patients and healthcare providers. This study aims to improve the diagnostic accuracy of resting ECG by employing deep learning techniques to remove motion artifacts, thereby demonstrating enhanced signal usability.

Recent advances in deep learning and neural networks have shown promise in addressing these challenges. By employing deep convolutional neural networks (CNN), it is possible to automate motion artifact removal in resting ECG signals and generate cleaner outputs for medical professionals to analyze. This study aims to develop a novel deep learning-based approach for motion artifact removal in resting ECG signals and verify its effectiveness by comparing it with existing methods.

## 2. Related Work

The challenge of motion artifact removal in ECG signals has been addressed through multiple signal processing and machine learning approaches. Early adaptive filtering methods [7] utilized custom motion sensors as reference inputs but demonstrated limited effectiveness for low-frequency noise components. Subsequent work employed Empirical Mode Decomposition (EMD) for real-time artifact detection in ambulatory monitoring systems [8], though without providing robust removal capabilities.

Wavelet-based techniques have shown particular promise, with Stationary Wavelet Transform (SWT) proving effective for artifact suppression while preserving QRS complex morphology [9]. Comparative studies have demonstrated SWT’s superiority over conventional Discrete Wavelet Transform (DWT) for ECG de-noising applications [10]. Independent Component Analysis (ICA) approaches [11] achieved noise separation but struggled with motion artifact specificity, particularly in 12-lead configurations.

Recent advances in deep learning have revolutionized ECG analysis. Personalized 1D CNNs have been developed for patient-specific ECG classification [12], while hybrid architectures combining wavelet transforms with neural networks have shown improved denoising performance [13]. Template-based adaptive filtering methods [14] and comprehensive noise removal frameworks [15] have further advanced the field.

Two-stage processing pipelines integrating wavelet denoising with machine learning classifiers [16] have demonstrated particular effectiveness. Deep learning approaches for automated ECG quality assessment [17] have achieved state-of-the-art results, though computational complexity remains a concern for real-time implementations. The current work builds upon these foundations while addressing key limitations in artifact removal completeness and computational efficiency.

### 2.1. Deep Learning & Attention Mechanisms

Recent developments have introduced novel deep learning methods like “AnEEG” for effective artifact removal from physiological signals, demonstrating improved quantitative metrics [18]. Advanced attention mechanisms, particularly the One-Dimensional Efficient Multi-Scale Attention Mechanism (1D-EMSAM), have emerged as powerful tools for extracting morphological and temporal features while separating physiological signals from artifacts [19]. These developments represent a paradigm shift from the primarily CNN-based approaches of the late 2010s, incorporating self-attention mechanisms that can capture long-range dependencies in ECG signals more effectively than traditional convolutions.

### 2.2. Multi-Modal and Multi-Resolution Approaches

Contemporary research has increasingly focused on multimodal fusion techniques that leverage complementary information sources for enhanced artifact removal performance. The development of multi-layer multi-resolution spatially pooled networks (MLMRS-Net), specifically for motion artifact removal in ambulatory EEG settings, has shown promising results [20], though similar approaches for ECG remain underexplored. Multi-modal image fusion with attention mechanisms has demonstrated significant improvements, with recent studies reporting F1-score improvements of 29%. Comprehensive reviews of motion artifacts in capacitive ECG monitoring systems have highlighted the need for more robust reduction techniques that can handle diverse noise sources in real-time monitoring applications, emphasizing the growing importance of multi-sensor approaches that combine accelerometer data with traditional ECG processing.

### 2.3. Transformer-Based Approaches and Hybrid Architectures

The integration of transformer architectures has revolutionized ECG signal processing and artifact removal capabilities. Constrained transformer networks embedded within CNN frameworks have demonstrated superior performance in capturing temporal information of ECG signals [21], while transformer-based deep neural networks like ECG DETR have shown remarkable success in continuous ECG signal processing [22]. Most notably, hybrid CNN-transformer models utilizing advanced time-frequency transforms like the Stockwell transform have achieved state-of-the-art performance in arrhythmia classification without requiring R-peak identification [23]. The emergence of CAT-Net (Convolution, Attention and Transformer based network) represents the latest evolution in single-lead ECG processing, demonstrating how multi-scale attention mechanisms can be effectively combined with transformer architectures for improved artifact detection and signal quality assessment [24].

### 2.4. Current Limitations and Future Directions

Despite significant advances, several critical gaps remain in the current state-of-the-art that limit clinical applicability and real-time deployment. While hybrid approaches combining recurrent neural networks with deep neural networks have shown promise for motion artifact removal, computational complexity remains a significant barrier for resource-constrained wearable devices. Recent surveys indicate that transformer and large language model applications to ECG diagnosis face challenges related to model interpretability and clinical validation [25, 26]. The field lacks standardized benchmarking protocols for comparing different artifact removal techniques and most current approaches focus on single-modality solutions rather than leveraging the multi-sensor capabilities of modern wearable devices. Furthermore, the majority of existing methods have been validated primarily on standard databases like MIT-BIH, with limited evaluation on diverse patient populations and real-world clinical conditions, highlighting the need for more comprehensive validation studies and standardized evaluation frameworks that can bridge the gap between research achievements and clinical implementation.

## 3. Methodology

### 3.1. Data Preparation and Collection

When a resting ECG is taken, an encrypted data file is created that comprises all lead information as well as important raw data. While a normal ECG setup includes a mechanism to convert such data to ASCII format, retrieving and interpreting such data is entirely dependent on the device manufacturer. As such, we considered the publicly accessible *PhysioNet* database [27] containing a 12-lead resting ECG. Prior knowledge included in these corpus enabled us to design proposed approaches capable of achieving the desired results in a laboratory context.

#### 3.1.1. Data Pre-processing

Surface measurements of the human body always contain a variety of noise that must be filtered out [15] before attempting to process the data using any pipelined technique. Baseline wandering is a term that refers to a sort of slow frequency noise in the ECG signal that results in a fluctuating baseline. The primary cause of wandering is the impedance between the ECG leads and the patient’s skin, which changes as the chest moves slightly while breathing, altering the distance between the heart and the leads. Although, baseline wandering may result in an incorrect ECG interpretations, such abnormalities can be estimated and removed [28, 29]. While these methods effectively address baseline wandering, they demonstrate limited efficacy in suppressing persistent motion and muscle artifacts.

### 3.2. Removal of Motion Artifacts

Visual interpretation of an ECG is challenging because motion artifacts are often mixed with raw ECG data to create an output that obscures each heartbeat 30. We will cover implementation specifics, tools utilized and the mathematical foundations for motion artifact reduction in the following sections.

#### 3.2.1. Methodology Selection

Because the primary objective of this research is to automate the removal of motion artifacts and to assess the processed signal using a machine learning model, the feasibility of implementing each technique is critical for reaching acceptable results. As in Section 2, the bulk of prior work is related to hardware implementations. In comparison, we offer a software solution that makes use of machine learning techniques to accomplish the primary goal.

#### 3.2.2. Implementation

It must be noted that only lead I data is considered in this section for the sake of explanation and clarity. We calculate the outlier ŝ _outliers_ and obtain a cleaned signal using equation 1, *n* ∈ [1, *N*] where *N* is the number of ECG samples as

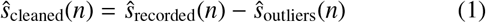

Calculating the amount of noise present in the signal is possible and can be used to obtain a cleaned ECG because when two signals overlap, the artifacts and signals are added up to produce the result by using SWT coefficient sequence and multiresolution thresholding, as depicted in Fig. 1.

**Figure 1.**
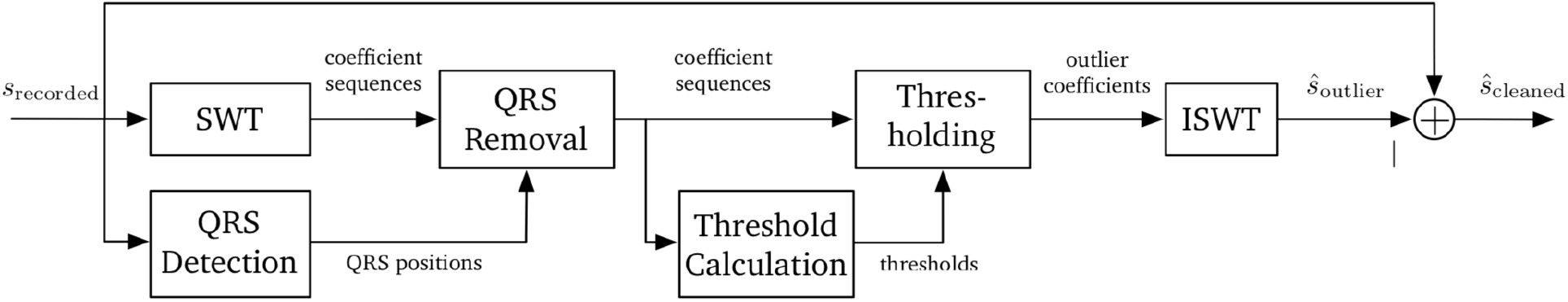
Schematic diagram of the multi-resolution thresholding algorithm [9].

Consider the role of DWT [31]. It decomposes a signal into a collection of mutually orthogonal wavelet functions which are dilated, translated and scaled copies of a common wavelet termed as a “mother wavelet”. Each DWT has two filters referred to as the *filter-bank* [31] which calculates the coefficients of a discrete collection of child wavelets for a given mother wavelet. Because DWT is applicable to non-stationary signals and the ECG meets this criterion, this technique becomes a well-established method for ECG de-noising and is capable of decomposing the signal to correct the relevant frequency spectrum. However, one of DWT’s primary disadvantages is its lack of translation invariancy. Assume there is a signal X and, even with a periodic signal extension, the DWT of a translated version of X is not, in general, the translated version of the DWT of X. This demonstrates the need of using SWT, which compensates for DWT’s lack of translation invariance by averaging a slightly modified DWT termed ε-decimated DWT.

The main two reasons for the selection of SWT [32] in this proposed work are as follows:

- SWT removes the translation-variance of the DWT because SWT is invariant against translations of the signal in the time domain [33].
- As the decimation of the coefficients at each level of the transformation is omitted, more samples in the coefficient sequences are available to detect possible outliers [9].

Implementing SWT is based on the method proposed in [34]. As depicted in Fig. 2, SWT generates two sequences, a wavelet coefficient and a scaling coefficient denoted by {*w*_1_, *w*_2_, …, *w*_*M*_ }and {*d*_1_, *d*_2_, …, *d*_*M*_,} respectively where *M* is the decomposition level of the SWT and *d*_0_ is the input signal. In this implementation, the very first *M* has to be calculated.

**Figure 2.**
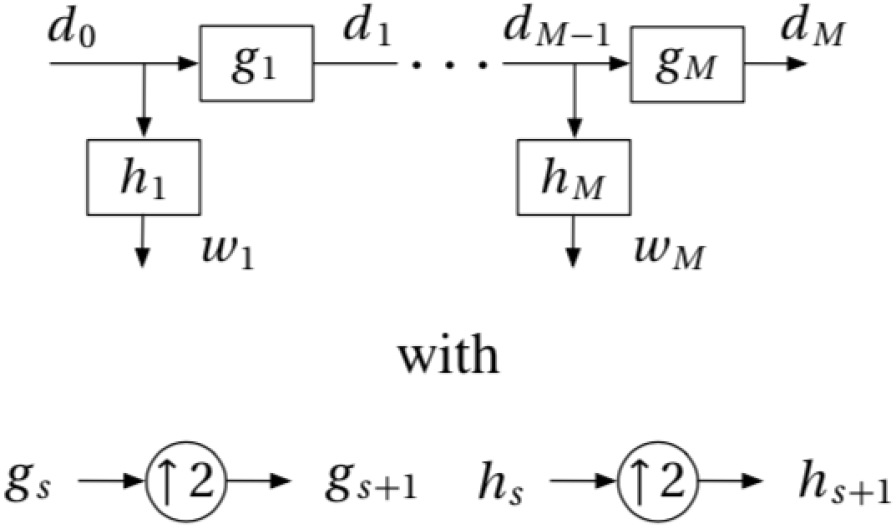
Implementation of SWT where h_s_ and g_s_ denote responses of low pass and high pass filters, respectively. The operator ↑ upsampling with a factor of 2 [34, 35].

De-noising an ECG signal involves several steps starting from SWT using the Haar as the mother wavelet as shown in Fig. 1. At each step, the relevant normalized median absolute deviation is used to compute a threshold. Finally, the threshold signals are converted back to ECG using an inverse SWT function. After de-noising the ECG [36], we proceed to identify the QRS complexes. For this purpose, the de-noised signal is subjected to a high pass filter to remove low frequency signals as shown in Fig. 3 [37].

**Figure 3.**
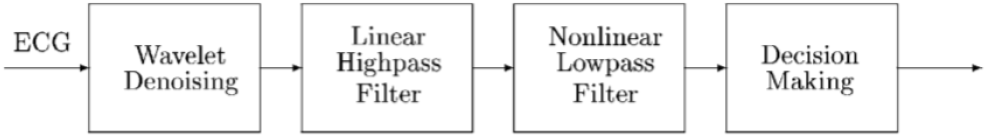
QRS detection pipeline [37].

According to Fig. 4, initially, the de-noised signal passes through an M-point moving-averaged linear high pass filter to accentuate the QRS complex. This filter with an ideal delayed system is used at this point as it causes the input function to shift forward in the time domain. The filtering process is followed by subtracting the result from the delayed input samples, point wise, to produce a high pass filter (HPF) having a finite impulse response (FIR). Let us consider *N* number of ECG samples where *n* ∈ [1, *N*]. Then, the output *y*_1_[*n*] and the delayed sample *y*_2_[*n*] are calculated as

**Figure 4.**
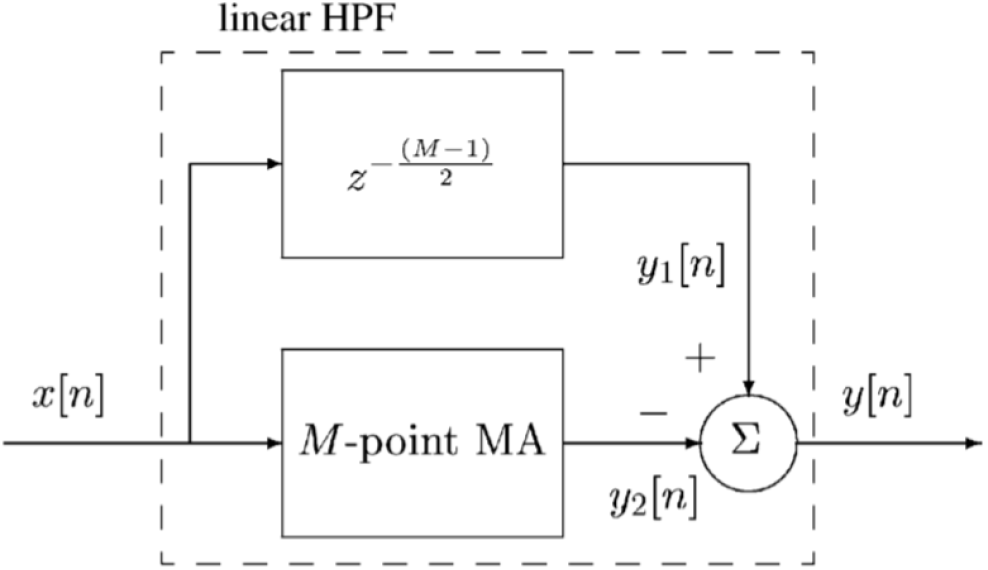
Architecture of HPF [37].

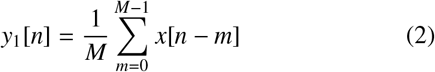

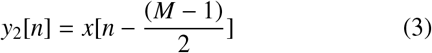

where *x*[*n*] represents the de-noised ECG input data and *M* is the filter length. If *M* is too large, P or T waves will be adversely enhanced [37]. As such, for convenience, *M* is restricted to an odd value heuristically initialized to 5. The output of the high pass filter is calculated as

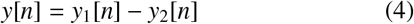

The next phase of the QRS detection mechanism is to send the output of the HPF into a non-linear low pass filter (LPF). This filtering technique is constructed using a cascade of point-by-point squaring operation having a moving window integration [37]. If *K* denotes the summation interval (or window length), the QRS feature *z*[*n*] is given as

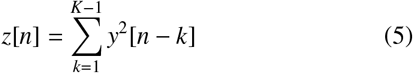

Because the threshold proposed by Chen et al.[9] is not robust against motion induced artifacts, a new threshold *T*_*QRS*_ is expressed as

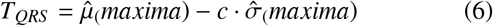

where 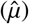 and 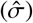 denote strong estimates of the mean and scale, respectively. Also the value for *c* has to be selected carefully because it depends on prior amount of impulsive noise in the data. The constant *c* should be chosen greater or equal to 2 to ensure detection of all QRS complexes and it depends on the prior knowledge of the amount of impulsive noise present in the data. In case impulsive noise is not included in the data, *c* is kept at above 2.5. However, as noise is inevitable in raw ECG, it is advisable to maintain *c* ∈ [2, 2.5]. As we have evaluated *T*_*QRS*_ from equation 6, the detected QRS, *QRS* _*Detection*_ is calculated as where

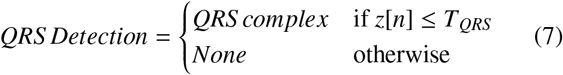

*Where QRS* _*complex*_ gives a valid QRS signal.

Prior to the last step in motion artifact reduction, a thresholding must be done following the sequence of processes as described above. To begin with, the lower and higher threshold sizes are determined using the SWT findings while the number of ECG data points and sequences for thresholding (SWT outputs) must be recorded. Subsequently, the upper and lower thresholds for each sequence are computed.

For this purpose, maxima and minima of estimated median 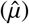 and scale 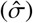 is used to calculate the least upper bound (*c*_*max*_) and the least lower bound (*c*_*min*_) as

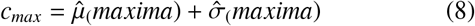

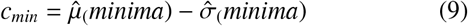

where 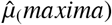 and 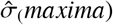 denote the maxima of the estimated median and scale, respectively. Also, 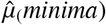 and 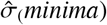 give the minima of estimated median and the scale, respectively. The upper threshold, *T*_*u*_, is then calculated as

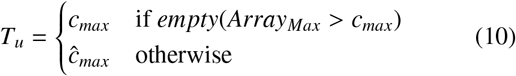

where *Array*_*Max*_ is the array containing the maximum value of each sequence and

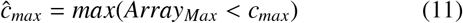

The lower threshold, *T*_*l*_, is obtained by

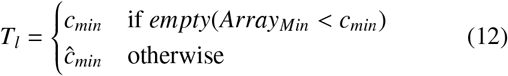

where *Array*_*Min*_ is the array containing the minimum value of each sequence and

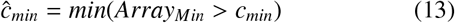

Subsequently, new wavelet coefficient sequences 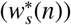 and scale coefficient sequences 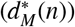 are calculated as

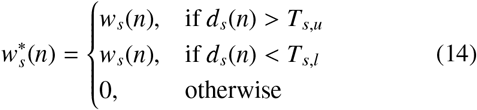

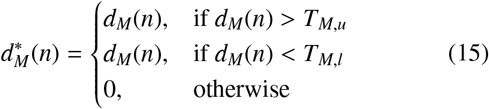

where *d*_*s*_(*n*) and *T*_*s,u*_ are the scale coefficient and upper threshold of level s, respectively while *T*_*s,l*_ gives the lower threshold of level s. Also, *T*_*M,u*_ and *T*_*M,l*_ denote the upper and lower thresholds of level M.

The method shown in Fig. 5 produces a perfect reconstruction of the signal, where 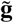 gives the inverse scaling filter coefficients and 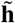 is the inverse wavelet filter coefficients.

**Figure 5.**
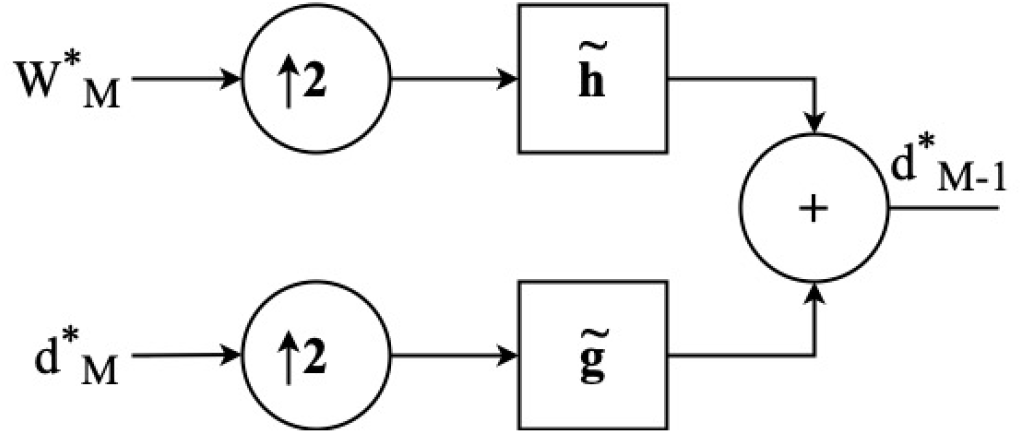
Steps of the Inverse Stationary Wavelet Transform (ISWT).

### 3.3. Deep ConvNet for ECG Evaluation

The 2^*nd*^ step of this study includes the implementation of a deep CNN to determine whether artifact-free ECG signals can be utilized for diagnostic purposes. Clinicians often identify any ECG abnormality using their professional knowledge accumulated throughout the years. Yet, a clinician may need to examine each wave in the stream in order to get a useful visual representation of the signal.

We utilized machine learning techniques to interpret ECG signals and compare the findings with ground truth acquired from an expert in order to verify whether the program will be able make predictions accurately and such predictions must be generated quickly.

#### 3.3.1. Data Preparation

Selecting an appropriate architecture and amassing relevant data for training, evaluation and testing are often difficult challenges when deploying CNNs. This section will discuss the methods used to gather and prepare data for this purpose.

As this study examines all 12 ECG leads, each lead may be assessed individually. Due to the fact that an ECG repeats the same signal (although not identical) at each beat and just a few seconds of the ECG are used for resting diagnosis, the size of each signal may be further reduced. Over 3000 image having a resolution of 74×448 pixels each are produced and saved in this experiment depending on the relevant leads. Additionally, to simplify training and testing, the data is randomly mixed via TensorFlow before feeding into the CNN. Each lead had three folders containing testing, training and validation data that are randomly split using the 80% − 20% separation method (80% for training and the remainder for validation).

#### 3.3.2. Architecture

It is standard practice in the research community to use a pretrained ConvNet model using millions of images classified into thousands of categories and to utilize it as a feature extractor or weight initializer for a particular job [38]. Finding a suitable network that is pre-trained for the area of interest, on the other hand, is challenging. The most difficult part of developing an architecture from scratch is assessment of performance heuristically which consumes a significant amount of both human and physical resources. Thus, transfer learning is used in a specific portion of this study and will be addressed in more detail later.

As we are unable to locate a pre-trained ConvNet in the ECG domain, an already-tuned network is modified as a significant quantity of data with augmentation is already available to train such a network. We utilized a simplified version of VGGNet [39] to detect noise in the ECG as such architectural modifications have been used in the literature to improve the accuracy in a variety of deep learning applications [40, 41]. This model comprises of four weight layers separated by ReLU and *Maxpooling* layers in which *Sigmoid* activation function is used to maximize the binary classification outcome in the final layer.

#### 3.3.3. Model Implementation

All model parameters, such as the batch size, number of epochs, etc., must be provided during model implementation. Table 1 indicates the number of images are collected for each lead. To produce a significant number of data for training, testing and validation, the images are enhanced using the Keras Image augmentation package. In addition, this library applies modifications with randomly chosen parameters within the given range to all the original pictures in the training set, guaranteeing that identical images are never generated at each learning epoch..

**Table 1:** Actual number of images used based on leads and categories.

| Leads | Number of images |  |
| --- | --- | --- |
|  | with noise | without noise |
| I | 1565 | 1565 |
| II | 1565 | 1565 |
| III | 1565 | 1565 |
| aVR | 1565 | 1565 |
| aVF | 1565 | 1565 |
| aVL | 1565 | 1565 |
| V1 | 1565 | 1565 |
| V2 | 1565 | 1565 |
| V3 | 1565 | 1565 |
| V4 | 1565 | 1565 |
| V5 | 1565 | 1565 |
| V6 | 1565 | 1565 |

**Table 2:** MSE values against M (decomposition level).

| M value | 1 | 2 | 3 | 4 | 5 | 6 | 7 | 8 | 9 | 10 |
| --- | --- | --- | --- | --- | --- | --- | --- | --- | --- | --- |
| MSE ($10^{-3}$) | 2.1 | 5.0 | 8.1 | 10.2 | 3.1 | 2.1 | 1.4 | 1.8 | 2.2 | 2.9 |

Moreover, the images had to be padded to make them square because the originals are of the size 74 × 448 pixels. Recent work has shown that padding images to create square shapes provide more accurate results than non-padded image data sets during training, testing and validation 39, 42, 43.

Another crucial fact to note is that the model may not be trained properly owing to variations in pixel intensities between training, validating and testing images. To resolve this issue, it is essential to treat all of these images uniformly. This crucial step is handled by *Rescale*, which configures the image rescaling factor where if 0 is given for *Rescale*, no rescaling occurs. However, when a valid scale is specified, the intensity value of each pixel in the image is multiplied by that scale prior to any image alteration to ascertain that the contribution of each image is identical and also to utilize the average pace of learning that we indicate.

As previously mentioned, some images include pixels with a high pixel density while others have pixels with a low pixel density. Without a scaling function, the model continues to use all such images with weights and learning rate. This is a major drawback and we will have an effect on its accuracy because when black and white data are evaluated, images with a higher white pixel count contribute more to the total loss than images with a lower white pixel count. However, back propagation will account for all such values lost without any preference to a particular image. Also, as a cleaned ECG has a higher percentage of white pixels, this tendency has the practical effect of decreasing the accuracy in binary classification. However, this effect may be significantly reduced by minimizing the contribution of white color.

#### 3.3.4. Training

Prior to the training step, we must select a suitable model for ECG assessment. This can be accomplished by using several models to evaluate the 12 leads or by using a single model to evaluate all 12 leads. Using several models for the evaluation is attempted by assigning the first model to *Lead I* according to the selected architecture for the training after augmenting 3, 000 images to 23, 500. This process is continued by augmentation 15, 000, 8, 500 and 3, 750 images for training, testing and validation, respectively.

After training a single model, the researcher utilizes transfer learning to create appropriate models for the other 11 leads using Keras and the previously trained model’s HDF5 data. The disadvantage of employing multiple models is that several models are required to detect motion artifacts in all 12 leads. However, resource allocation may be reduced significantly if a single model is employed to detect such artifacts in all 12 leads. To do this, the data from the 12 leads is divided into two categories: with and without noise. Following that, the 36, 000 pictures are increased to 47, 000 and divided into 30, 000, 17, 000 and 7, 500 images for training, testing and validation, respectively.

## 4. Results and Discussion

### 4.1. Motion Artifacts Removal

The results of a single lead (Lead I) ECG data are presented in this section for the sake of illustration.

Fig. 6 illustrates an ECG that is generated for validation purposes and has obvious motion artifacts, most notably, around the fourth heart beat. As a consequence, this may be used to determine if the chosen method performs as anticipated.

**Figure 6.**
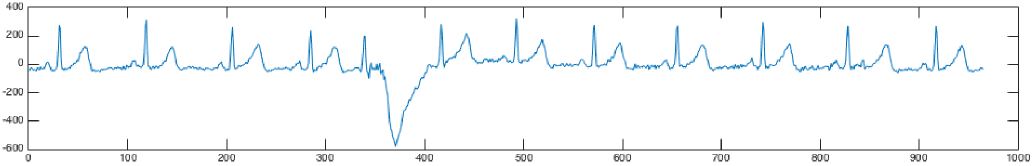
The ECG generated by data augmentation methods similar to ECG used in [9].

As shown in Fig. 7, each level of the SWT is detected and utilized to eliminate relevant noise components from the original signal using five wavelet coefficients. This enables us to prevent QRS detection by highlighting outliers that can be eliminated.

**Figure 7.**
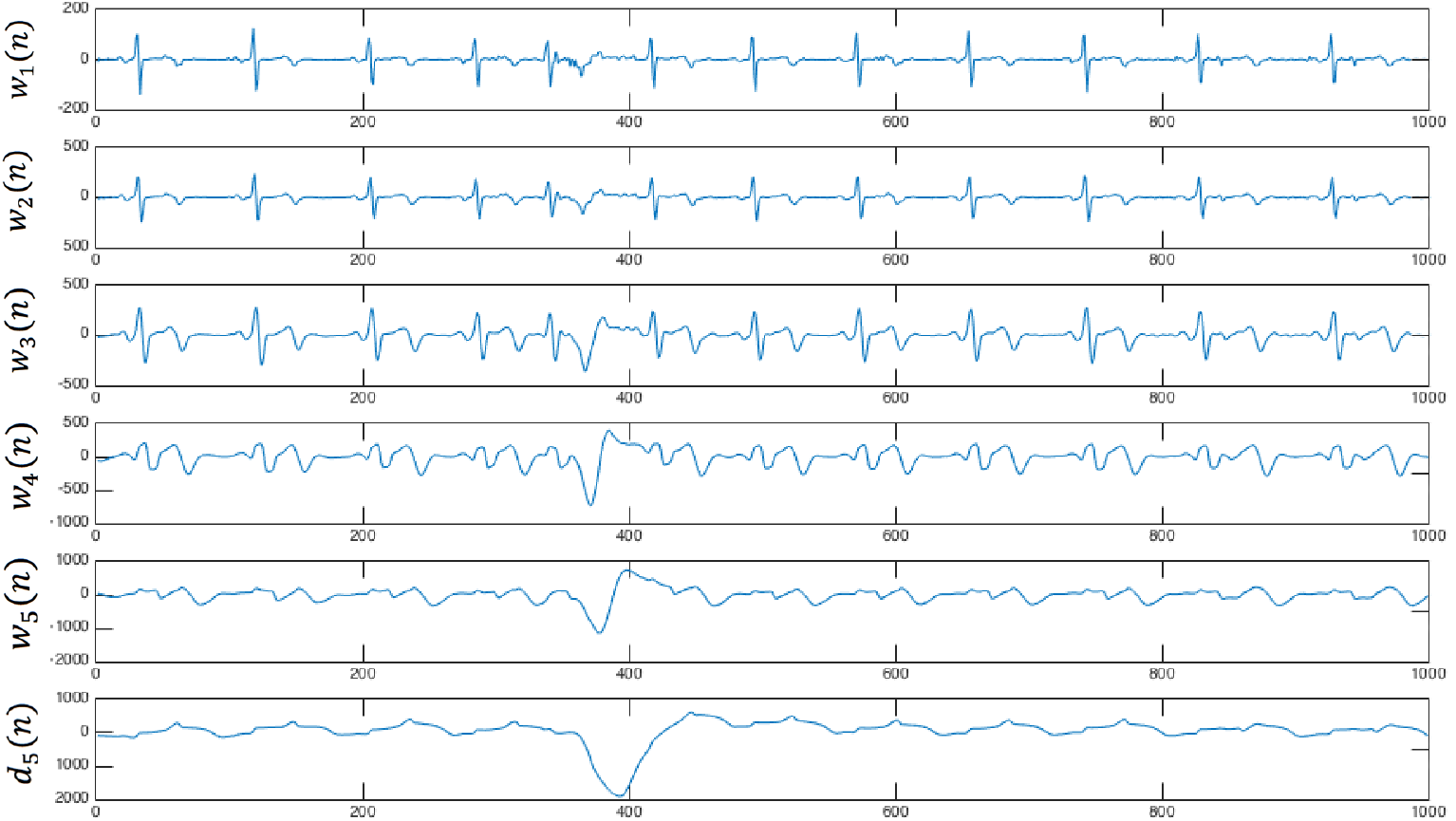
Decomposition levels after applying SWT on the ECG.

Fig. 8 depicts the thresholds of the ECG obtained to calculate the error. To distinguish outliers from the actual ECG signal, we compute an upper and lower thresholds for each coefficient sequence by dividing each sequence into segments as explained in Section 3 and subsequently, the maxima and minima for each sequence is computing using Equations (8, 9).

**Figure 8.**
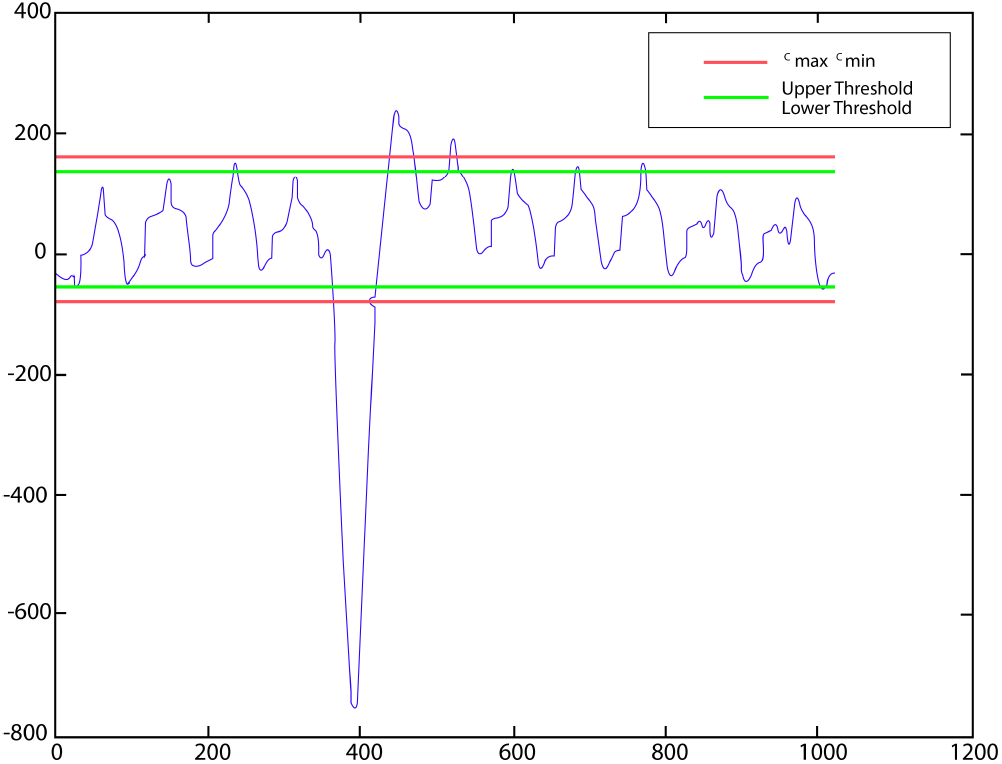
Thresholds with bounds.

Fig. 9 clearly shows the error calculated from raw ECG data. Although this error is deducted from raw data to produce a corrected ECG, the it may not exhibit 100% accuracy. To address this issue, *Savitzky-Golay filter* [44] is introduced to improve the signal clarity while preserving the accuracy.

**Figure 9.**
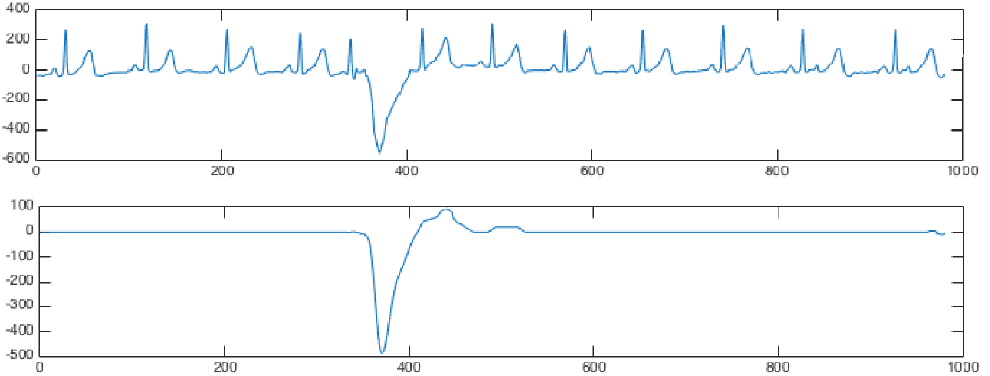
Raw ECG data (top) and error (bottom).

Fig.10 shows the effect of filtering ECG signals where we analyzed 500 unprocessed 1000Hz-generated ECGs from healthy and unhealthy subjects which produced an MSE = 0.0014 with filtering and MSE = 0.0018, without filtering. Moreover, we set M = 7 in this interpretation as it produced the lowest MSE Table Fig 11 presents the processed ECG signal following motion artifact removal and smoothing, demonstrating improved signal quality.

**Figure 10.**
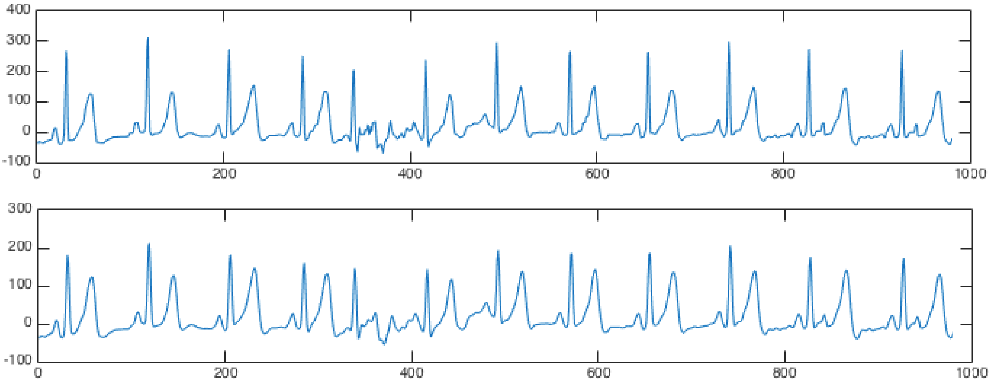
ECG signals (top) without filtration and (bottom) with filtration.

**Figure 11.**
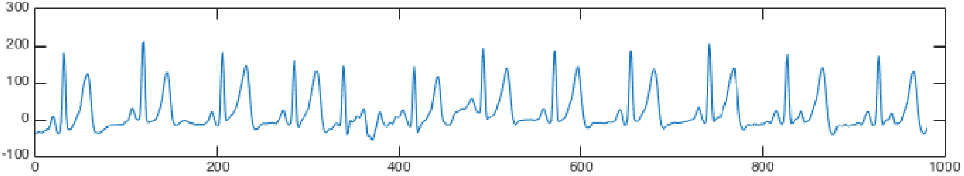
Lead I ECG after removing motion artifacts and improving details with Savitzky-Golay filter.

### 4.2. Deep ConvNet for ECG Evaluation

This section explains all intermediate and the final outputs of ConvNet training.

#### 4.2.1. Data Preparation

Data preparation is one of the most time-consuming activities in this stage because a huge data collection is required to commence the task. A single ECG signal may not usually provide sufficient information to remove noise components due to rapid movements of the subject resulting in noise together with increased heart rate. Thus, an ECG with two or five beats is insufficient to detect the presence of significant noise when the time laps is short. To avoid this barrier, we created a method for capturing pictures in an ECG with ten to fifteen beats as seen in Fig. 12.

**Figure 12.**
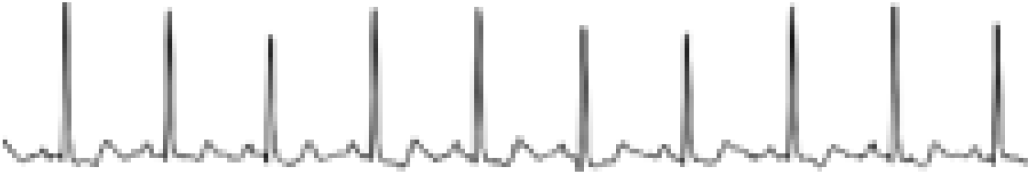
Sample lead I ECG image used to train the models (without noise).

#### 4.2.2. Models Evaluation

Determining the optimal number of convolutional neural networks (ConvNets) required to achieve high classification accuracy presents significant computational challenges. As detailed in Section 3, we address this through two distinct approaches: (1) training individual models for each of the 12 ECG leads and (2) developing a unified model encompassing all leads. To optimize input processing, we systematically evaluated image padding techniques by comparing model performance with and without padding. Our experiments demonstrated superior accuracy for padded inputs (98-99%) compared to unpadded alternatives (95.8-96%), establishing padding as the preferred preprocessing method for the final architecture.

The Table 3 shows the training accuracy of each model trained to evaluate relevant lead data.

**Table 3:** Performance of trained models.

| Model<br>(lead specific) | TP | TN | FP | FN | Sen.% | Spec.% | Prec.% | Training<br>accuracy % |
| --- | --- | --- | --- | --- | --- | --- | --- | --- |
| I | 315 | 309 | 9 | 3 | 99.06 | 97.17 | 97.22 | 98.11 |
| II | 288 | 318 | 3 | 32 | 90.00 | 99.06 | 98.97 | 94.54 |
| III | 315 | 314 | 4 | 5 | 98.44 | 98.74% | 98.75 | 98.59 |
| aVR | 314 | 320 | 0 | 6 | 98.13 | 100.00 | 100.00 | 99.06 |
| aVF | 313 | 315 | 5 | 6 | 98.12 | 98.44% | 98.43 | 98.28 |
| aVL | 309 | 316 | 4 | 11 | 96.56 | 98.75 | 98.72 | 97.66 |
| V1 | 314 | 315 | 5 | 6 | 98.13 | 98.44 | 98.43 | 98.28 |
| V2 | 314 | 318 | 2 | 6 | 98.13 | 99.38 | 99.37 | 98.75 |
| V3 | 290 | 319 | 1 | 30 | 90.62 | 99.69 | 99.66 | 95.16 |
| V4 | 309 | 316 | 4 | 11 | 96.56 | 98.75 | 98.72 | 97.66 |
| V5 | 315 | 316 | 4 | 5 | 98.44 | 98.75 | 98.75 | 98.59 |
| V6 | 297 | 318 | 2 | 23 | 92.81 | 99.38 | 99.33 | 96.09 |

After training the single model to assess data taken from the 12 leads, we achieved an overall accuracy of 98 percent. This overarching figure is strongly supported by the granular data from the individual leads, which reveals the robustness of the approach. The strategy of training separate models for each lead has paid significant dividends, as it provides a nuanced diagnostic capability that a single, monolithic model could not offer.

The consistency of high performance across most leads is a key indicator of a reliable system. Leads such as aVR, which achieved a perfect specificity (100%) and precision (100%), along with V2 and III, show that the model is exceptionally adept at correctly identifying true negatives and that its positive predictions are incredibly trustworthy. This is of paramount importance in a clinical setting, where minimizing false alarms (FP) is crucial to avoid unnecessary patient anxiety and additional testing. Furthermore, the very high sensitivity scores, many above 98%, mean the model is also highly effective at correctly identifying true positives, ensuring that genuine cases are unlikely to be missed.

However, the data also offers valuable insights into potential areas for investigation. The slightly lower performance of leads II, V3, and V6, primarily driven by a higher number of false negatives (FN), is not necessarily a weakness but a useful characteristic. It suggests that the cardiac information captured in these specific leads might be more ambiguous or varied for the condition the model is designed to detect. This doesn’t detract from the overall success; rather, it provides a clear roadmap for future work. Focusing on why these leads are more challenging could lead to an even more robust model, perhaps by enriching the training dataset with more examples of the variations present in these leads or by further refining the feature extraction process for them.

However, for each individual lead, Table 4 indicates the degree to which this assessment is correct.

**Table 4:** Performance of the single model trained to denote all leads.

| Leads | TP | TN | FP | FN | Sen.% | Spec.% | Prec.% | Accuracy % |
| --- | --- | --- | --- | --- | --- | --- | --- | --- |
| I | 317 | 319 | 1 | 7 | 97.84 | 99.69 | 99.69 | 98.76 |
| II | 317 | 318 | 2 | 3 | 99.06 | 99.38 | 99.37 | 99.22 |
| III | 313 | 318 | 2 | 7 | 97.81 | 99.38 | 99.36 | 98.59 |
| aVR | 318 | 320 | 0 | 2 | 99.38 | 100.00 | 100.00 | 99.69 |
| aVF | 311 | 319 | 1 | 9 | 97.19 | 99.69 | 99.68 | 98.44 |
| aVL | 308 | 317 | 3 | 12 | 96.25 | 99.06 | 99.03 | 97.66 |
| V1 | 316 | 315 | 5 | 4 | 98.75 | 98.44 | 98.44 | 98.59 |
| V2 | 319 | 314 | 6 | 1 | 99.69 | 98.13 | 98.15 | 98.91 |
| V3 | 319 | 316 | 4 | 1 | 99.69 | 98.75 | 98.76 | 99.22 |
| V4 | 320 | 310 | 10 | 0 | 100.00 | 96.88 | 96.97 | 98.44 |
| V5 | 319 | 315 | 5 | 1 | 99.69 | 98.44 | 98.46 | 99.06 |
| V6 | 319 | 318 | 2 | 1 | 99.69 | 99.38 | 99.38 | 99.53 |

The unified model architecture demonstrates significant computational advantages over the lead-specific approach. While the combined model requires only a single load operation before prediction, the 12-model alternative necessitates loading all individual networks prior to inference. Our benchmarking tests revealed a substantial performance difference: the unified model achieved prediction times of 1.60 ± 0.05 seconds (mean ± SD), compared to 21.67 ± 0.12 seconds for the multi-model approach. These metrics are derived from 100 repeated trials of model loading and evaluation, with timing data collected for each iteration and reported as mean values.

The results shown in Fig. 13 are evaluated by the selected model and subsequently confirmed by experienced clinicians. Fig. 14 illustrates an ECG with significant noise even after motion artifacts are removed hampering accurate visual diagnostics. This prediction is clearly made by the trained model within 2 seconds whereas an expert may take approximately 5 seconds for visual interpretations. In this scenario, the complete ECGs will be produced only if the physician selects the option to generate a full ECG.

**Figure 13.**
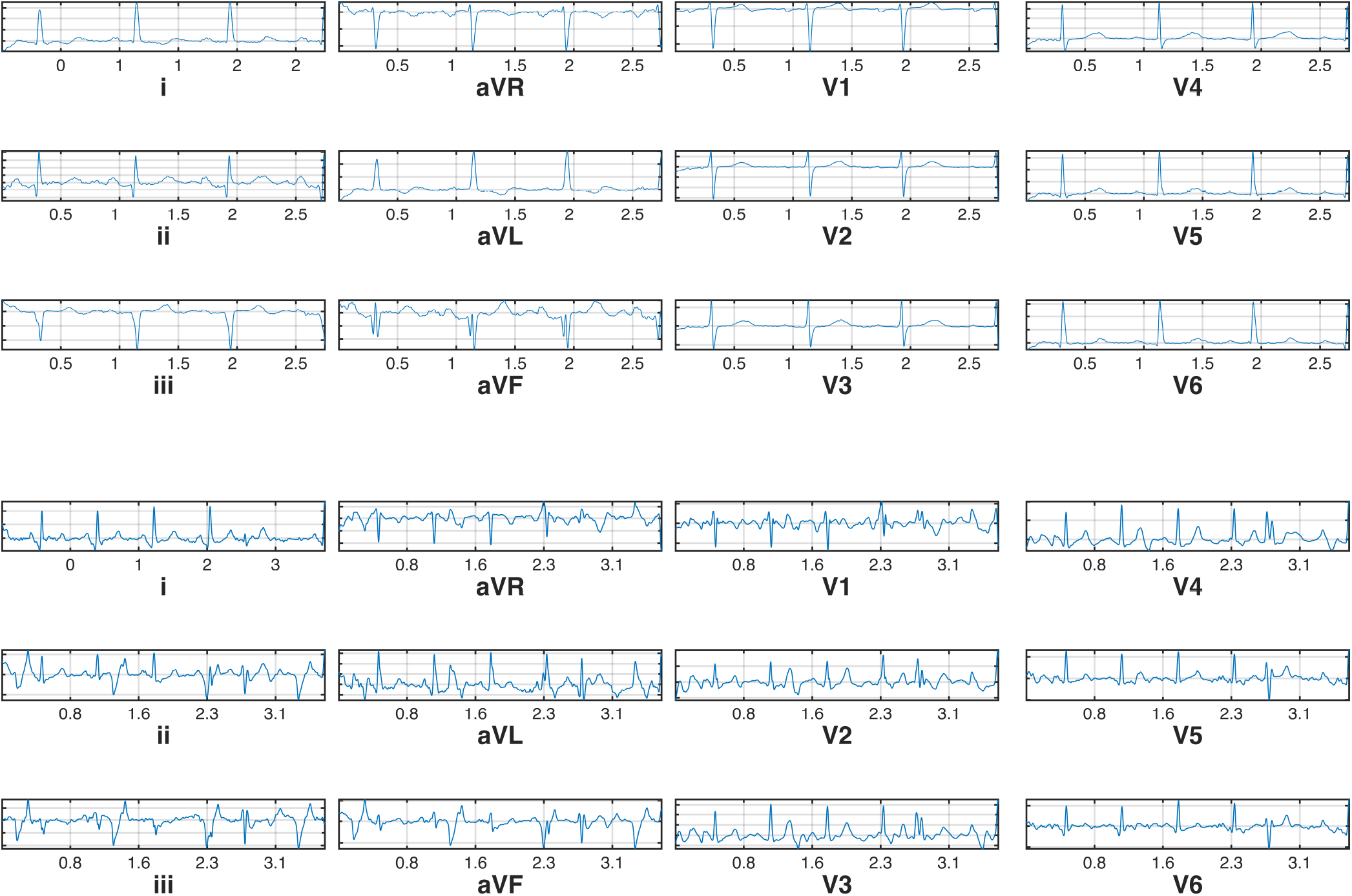
ECG without motion artifacts: (top) healthy and (bottom) non-healthy subjects.

**Figure 14.**
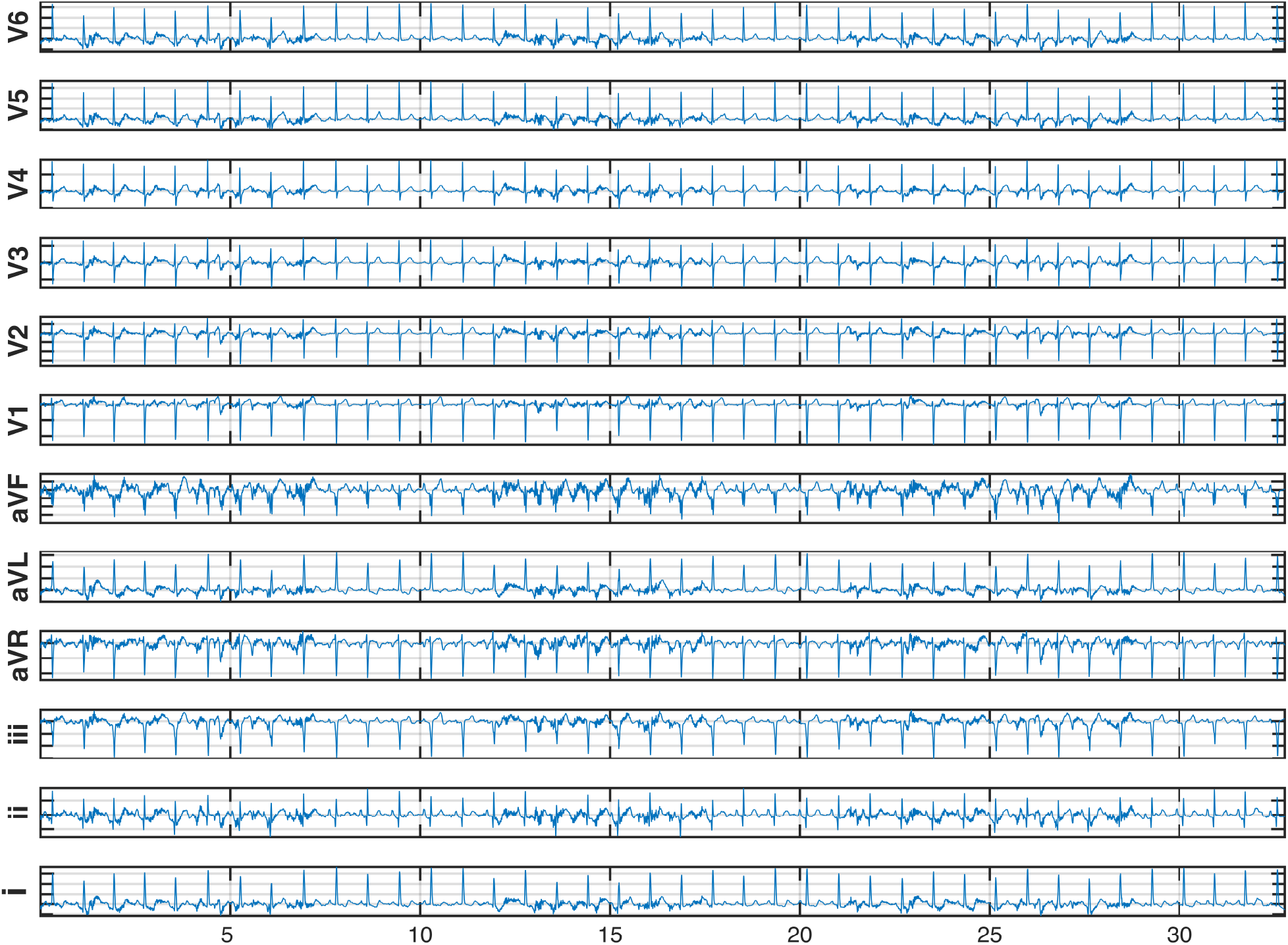
Full ECG found with heavy motion artifacts.

### 4.3. Motion Artifacts Removal

One of the most challenging aspects of this study is to reduce motion artifacts while retaining the correct ECG data that may possibly aid in identifying various underlying medical conditions. For this purpose, the assistance of many cardiographers are obtained to fine-tune application settings to produce an accurate output while ensuring that critical medical interpretation from the original ECG are not excluded.

As seen in Fig. 10, we may unequivocally determine that the *Savitzky-Golay filter* has enhanced the clarity of the ECG output. The MSE, on the other hand, must be computed to see if this filter had actually reduced this error and at the same time, making it more legible.

During the initial stage in eliminating motion artifacts, the M value, which represents the SWT’s decomposition level, is chosen to be 5. Initially, this value is chosen as explained in the [45] to obtain a better de-noising effect. Subsequently, after the routine is completed, another test is conducted to determine the SWT’s optimal decomposition level as in and Fig. 2. As a consequence, it is recommended to utilize M = 7 for the ECGs in the 1000Hz frequency range. This enabled us to reorganize the software to provide a highly accurate ECG output when motion artifacts are removed. The outcome of each procedure is evaluated by experts in the subject matter to fine-tune the software to provide the most accurate information possible.

### 4.4. Deep ConvNet for ECG Evaluation

An important task in this research is to determine the optimal number of machine learning models to train the data with. Initially, a single model is assigned for each lead which produced the results as given in Table 3 resulting in more than 95% of accuracy at all times. Next, a model is created using all leads combined or simply saying, a combined model, to evaluate the data which produced more than 97.66% accuracy at all times as shown in Table 4.

These result clearly demonstrate that when all leads are merged together, the combined model outperforms all other models except in Lead III and as such, it is preferable to utilize a mixed model for ECG assessment. The improved accuracy of the combined model may be due to each signal’s similarity when compared with one another as these signals only indicate the amount of voltage received by each electrode between the heart beats. As the lag time between the leads is negligible, certain ECG signals are not identical but somewhat comparable in signal strength. Hence, having over 30,000 images with such a resemblance will certainly assist the model in learning about signals and associated noise levels.

The unified model offers significant advantages in resource efficiency compared to training individual lead-specific models. Specifically, it reduces storage requirements by an order of magnitude (1 GB vs. 15 GB for 12 separate models) and accelerates inference time. This performance improvement stems from eliminating redundant operations: while the combined model loads parameters once, the multi-model approach incurs repeated I/O overhead by loading 12 independent H5 files sequentially.

### 4.5. Comparison with Multi-Task Learning (MTL) Approaches

Our unified model approach, which trains a single CNN to perform quality assessment across all 12 ECG leads, shares the high-level objective of multi-task learning (MTL) methods: to improve efficiency and generalization by leveraging commonalities between related tasks. However, it differs fundamentally in its architecture and learning mechanism from typical deep MTL frameworks.

Our approach can be viewed as a specific, highly efficient instantiation of the MTL philosophy. Instead of formalizing the processing of each lead as a separate task, we recast the problem into a single task with multiple, semantically similar inputs. This is a valid and powerful strategy because the “task”, assessing signal quality based on morphological features and noise patterns, is conceptually identical across all 12 leads. The leads are simply different electrical viewpoints of the same underlying cardiac activity. The main features of the proposed architecture and the deep MTL method are summarized in Table 5.

**Table 5:**
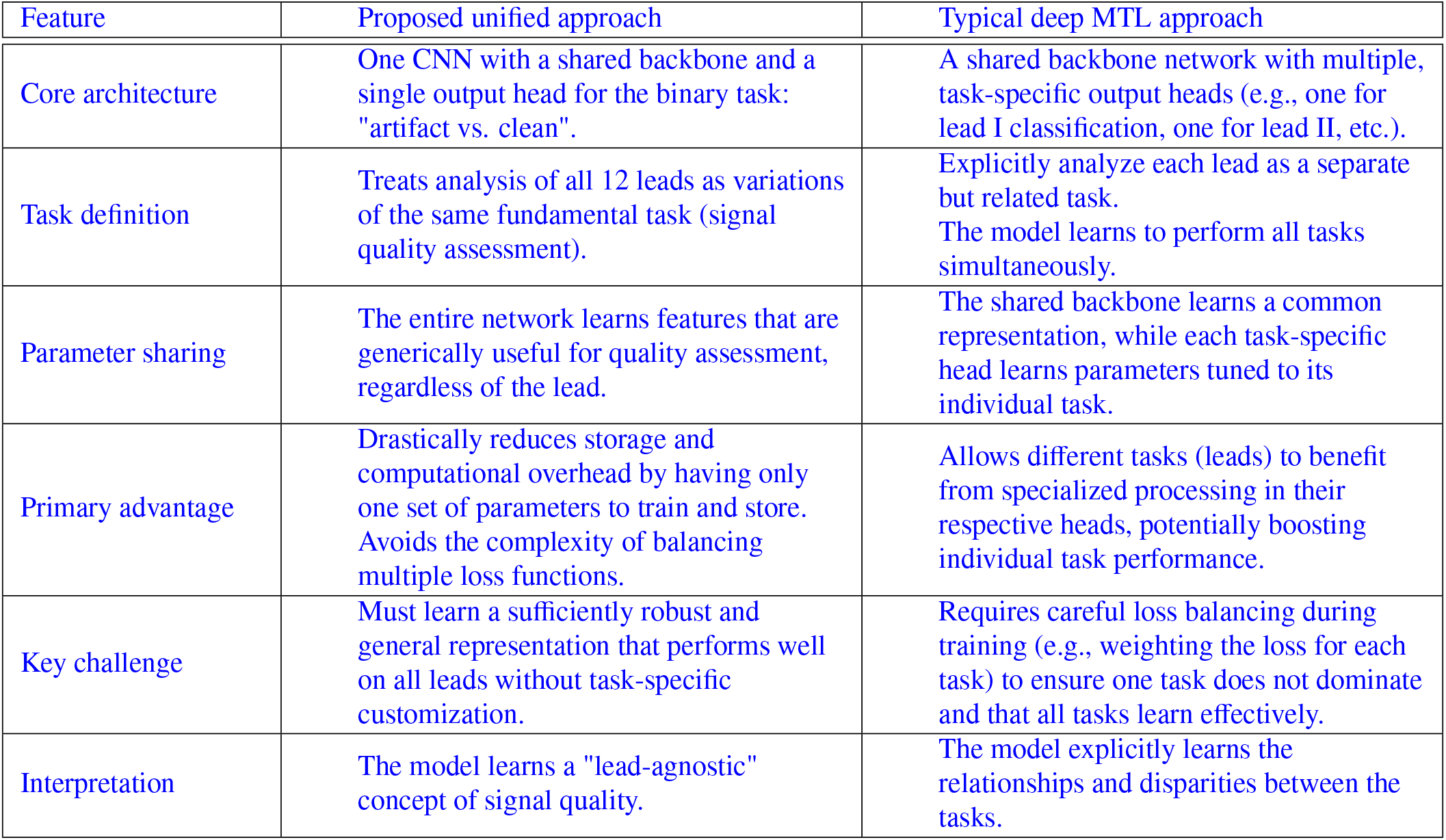
Comparison of proposed architecture with Deep MTL method.

| Feature | Proposed unified approach | Typical deep MTL approach |
| --- | --- | --- |
| Core architecture | One CNN with a shared backbone and a single output head for the binary task: "artifact vs. clean". | A shared backbone network with multiple, task-specific output heads (e.g., one for lead I classification, one for lead II, etc.). |
| Task definition | Treats analysis of all 12 leads as variations of the same fundamental task (signal quality assessment). | Explicitly analyze each lead as a separate but related task.<br>The model learns to perform all tasks simultaneously. |
| Parameter sharing | The entire network learns features that are generically useful for quality assessment, regardless of the lead. | The shared backbone learns a common representation, while each task-specific head learns parameters tuned to its individual task. |
| Primary advantage | Drastically reduces storage and computational overhead by having only one set of parameters to train and store. Avoids the complexity of balancing multiple loss functions. | Allows different tasks (leads) to benefit from specialized processing in their respective heads, potentially boosting individual task performance. |
| Key challenge | Must learn a sufficiently robust and general representation that performs well on all leads without task-specific customization. | Requires careful loss balancing during training (e.g., weighting the loss for each task) to ensure one task does not dominate and that all tasks learn effectively. |
| Interpretation | The model learns a "lead-agnostic" concept of signal quality. | The model explicitly learns the relationships and disparities between the tasks. |

**Table 6:** Comparison of proposed method with recent work.

| Feature | Proposed method | 1D-CNN [12] | Transformers [23] | CAT-Net [24] |
| --- | --- | --- | --- | --- |
| Primary objective | Artifact removal + quality assessment | Arrhythmia classification | Arrhythmia detection | Arrhythmia classification |
| Core approach | Two-stage: SWT-SG filter + Unified CNN classifier | 1D personalized CNN | Hybrid CNN-Transformer + Stockwell transform | Multi-scale convolution, attention & transformer |
| Input modality | 12-lead ECG (image plots) | 1D ECG signal | 1D ECG signal | Single-lead ECG signal |
| Key innovation | Unified model for all leads, efficient resource usage, integrated filtering | Patient-specific adaptation | Transformer for long-range dependencies without R-peak detection | Integrated multi-scale attention and transformer blocks |
| Artifact handling | Explicit removal & classification | Not a primary focus | Not a primary focus | Not a primary focus |
| Model architecture | Custom 2D CNN (VGG-based) | 1D CNN | CNN + Transformer encoder | CNN + attention + transformer |
| Computational efficiency | High (inference: 1.6s, storage: 1GB) | High (for 1D signal) | Low (high complexity from transformer) | Low (high complexity from transformer/attention) |
| Interpretability | Moderate (feature maps can be visualized) | Low (black-box 1D CNN) | Very low (complex self-attention maps) | Very low (complex multi-scale attention) |
| Data efficiency | High (uses spatial features from 2D plots) | Moderate (requires patient-specific data) | Low (transformers require large datasets) | Low (complex models require large datasets) |
| Clinical deployment | Highly suitable (fast, lightweight, automated QA) | Suitable for personalized monitoring | Computationally challenging for edge devices | Computationally challenging for edge devices |
| Reported performance | Accuracy > 98% in artifact reduction | High patient-specific accuracy | High arrhythmia detection accuracy | High classification accuracy |

This recasting allows us to capture the core benefit of MTL, learning a generalized representation that improves performance and efficiency, while avoiding its primary engineering complexity: the need to balance multiple loss functions. Our results demonstrate that this simplified approach is not only sufficient but highly effective, achieving excellent accuracy across all leads without any task-specific weighting or customization.

In contrast, more complex MTL architectures, such as those employing cross-stitch networks or uncertainty weighting, are typically justified when the tasks are more divergent (e.g., simultaneously detecting arrhythmias, estimating ejection fraction, and assessing signal quality). For the focused objective of lead-agnostic quality assessment, our unified model provides a more parsimonious and deployable solution.

While sophisticated MTL methods offer a powerful framework for learning multiple disparate tasks, our proposed unified model demonstrates that for closely related tasks like multi-lead ECG quality assessment, a simpler single-task, multi-input architecture can achieve superior computational and storage efficiency without compromising on classification accuracy. This makes it particularly suitable for resource-constrained clinical environments where model size and inference speed are critical.

### 4.6. Comprehensive Comparison of ECG Artifact Handling Methods

The Table 4.7 highlights the unique positioning and contributions of our work compared with other techniques. While recent state-of-the-art studies focus on end-task diagnosis like arrhythmia detection [23, 24], our work addresses a fundamental prerequisite: ensuring signal quality. A high-performing arrhythmia detector will fail on a corrupted signal. Our work provides a crucial “quality gate” that can be placed before these advanced diagnostic models. This clearly shows the trade-off between performance and efficiency. The proposed unified 2D CNN model sacrifices some of the theoretical representational power of transformers for superior computational and storage efficiency. This makes it far more suitable for real-time or resource-constrained clinical environments, which is a significant practical contribution. Further, unlike earlier two-stage methods that only classify quality [12], the proposed pipeline includes an advanced artifact removal stage (SWT + SG Filter), actively improving the signal, not just flagging it.

Our demonstration that a single model can effectively handle all 12 leads is a key innovation in efficiency, contrasting with the implicit assumption that complex, specialized models are always needed for high performance. This comparison ensures that our proposed framework does not directly compete with the latest diagnostic transformers but rather complements them. By providing a highly efficient, accurate and dedicated solution for the critical problem of motion artifact management, our work enables the reliable deployment of those more complex models in real-world clinical settings. The choice of a unified CNN architecture is justified by the need for speed, low resource consumption and high reliability, filling a distinct and vital niche in the ECG processing pipeline.

### 4.7. Novelty, Contribution and Scope

#### Novelty

While transformer-based architectures have demonstrated state-of-the-art performance in various sequence processing tasks, including recent applications in ECG analysis [23, 22], our choice of a CNN for the artifact classification stage was driven by several critical factors aligned with the specific constraints and objectives of this study. Transformers are notoriously data-hungry, requiring massive datasets to learn effective representations due to their large number of parameters and self-attention mechanisms. Our dataset, while substantial for a CNN-based image classification task (∼47,000 augmented images), is orders of magnitude smaller than the datasets typically used to train effective transformers from scratch. CNNs, with their strong inductive biases for spatial locality and translation invariance, are far more data-efficient and thus less prone to overfitting on our dataset size.

CNNs are a mature, well-understood technology with stable training dynamics and established best practices. This reduces experimental complexity and risk. Furthermore, the feature maps learned by CNNs can be more straightforward to visualize and interpret (e.g., via Grad-CAM) to understand which regions of the ECG image most influenced the classification decision, which is a valuable asset for clinical validation.

This work introduces several novel aspects to the field of ECG motion artifact detection and removal. Unlike previous approaches that focus solely on either artifact removal or quality assessment, this study presents an integrated framework that combines SWT preprocessing with deep CNN-based quality evaluation in a unified pipeline. Rather than training separate models for each of the 12 ECG leads, this work demonstrates that a single combined CNN model can effectively evaluate all leads simultaneously, representing a departure from traditional lead-specific approaches. The study proposes a modified threshold calculation that improves robustness against motion-induced artifacts compared to existing methods, specifically addressing limitations in [37]. Additionally, the research demonstrates significant computational efficiency improvements (1.6 vs. 21.7 seconds) while maintaining high accuracy, making the approach viable for clinical real-time applications.

Our core novelty lies in the integration of a sophisticated, wavelet-based preprocessing stage for artifact removal with a deep learning-based quality gate. The CNN serves as a highly effective and efficient classifier for this integrated pipeline. Investing complexity in the hybrid signal-processing/learning approach for artifact removal (Stage 1) and using a robust, efficient classifier (Stage 2) was deemed a more optimal design choice than attempting to force a single, complex transformer model to learn both artifact removal and classification simultaneously from limited data.

Our selection of a CNN architecture was a deliberate and strategic decision based on data constraints, computational efficiency goals, optimal alignment with the spatial nature of the classification task and the overall design philosophy of our twostage pipeline. While transformers represent a powerful emerging trend, the CNN provided the best balance of performance, efficiency and reliability for the specific objectives of this study. Future work, involving larger multi-modal datasets, could explore hybrid CNN-Transformer architectures to leverage the strengths of both paradigms.

## Contribution

The primary contributions of this research to the biomedical engineering field encompass both technical and clinical advances. From a technical perspective, this work achieves 98.76% classification accuracy in distinguishing between usable and artifact-corrupted ECG signals, with sensitivity of 98.74% and specificity of 98.77%. The study develops a resource-efficient solution that reduces storage requirements from 15GB (12 separate models) to 1GB (unified model) while accelerating inference time by approximately 13-fold. The methodological innovation integrates multi-resolution thresholding with Savitzky-Golay filtering to preserve critical ECG morphological features while effectively removing motion artifacts, supported by comprehensive evaluation using 12-lead ECG data with expert validation. From a clinical standpoint, the enhanced ECG signal quality reduces the risk of misdiagnosis caused by motion artifacts in resting ECG recordings. The automated quality assessment can reduce clinician workload by providing rapid (≤ 2 seconds) signal evaluation compared to manual interpretation, while offering a systematic approach to artifact detection that could improve consistency in ECG quality assessment across different clinical settings.

## Limitations

While this study presents significant advances, several limitations must be acknowledged across technical, methodological and clinical domains. The research relies on publicly available PhysioNet datasets, which may not fully represent the diversity of motion artifacts encountered in real clinical environments with different patient populations, recording conditions and equipment variations. The focus on motion artifacts means the system may not effectively handle other common ECG noise sources such as power line interference, electrode contact issues or electromagnetic interference from medical devices. The optimal decomposition level (M=7) was determined specifically for 1000Hz ECG signals, potentially limiting appli-∈ cability to systems using different sampling frequencies commonly found in clinical practice, while the study depends on expert visual assessment for ground truth labeling, introducing potential subjectivity and inter-observer variability. The current system has not been extensively tested across a comprehensive range of cardiac pathologies, which may affect its generalizability to diverse patient populations with various heart conditions. The thresholding parameters (c [2, 2.5]) are empirically determined and may require adjustment for different patient populations or recording conditions. Expert validation was conducted with a limited number of clinicians, requiring broader clinical validation studies to establish inter-rater reliability and clinical utility. The study does not address regulatory approval requirements or clinical validation standards necessary for deployment in healthcare settings, nor does it characterize system performance on borderline cases where artifact presence is ambiguous. Limited evaluation across different patient demographics, body types and clinical conditions that may affect ECG signal characteristics and artifact patterns represents another constraint that future research must address through expanded datasets, multimodal physiological signal incorporation, adaptive parameter selection mechanisms, largescale clinical validation studies and investigation of real-time integration capabilities with commercial ECG systems.

## 5. Conclusions

This study introduces a deep learning–based framework for the detection and removal of motion artifacts in resting ECG signals, aiming to improve diagnostic reliability in clinical settings. By integrating advanced preprocessing methods with a robust classification model, the proposed approach effectively distinguishes between clean and artifact-contaminated signals, thereby enhancing ECG usability for medical decision-making. The results demonstrate notable improvements in clarity and diagnostic quality, indicating that deep learning can serve as a valuable tool in minimizing the risk of misdiagnosis caused by motion-induced noise. Despite these achievements, the current system relies on manually collected ECG datasets and does not encompass all known cardiac disorders, which may limit its generalizability. Future work will focus on integrating the algorithm directly into ECG acquisition devices to enable real-time artifact detection and correction. Additionally, expanding the training dataset to include a broader spectrum of cardiac pathologies will improve robustness and reduce the likelihood of misclassification. Incorporating multimodal physiological signals, such as photoplethysmography (PPG) or accelerometer data, could further enhance the model’s artifact detection accuracy. Ultimately, these developments will advance the reliability, automation and clinical adoption of ECG-based diagnostics.

## Data Availability

The data underlying the results presented in the study are available from PhysioNet.

https://physionet.org/content/nstdb/1.0.0/

## Data Availability

The raw data required to reproduce the above findings are available to download from https://physionet.org/content/nstdb/1.0.0/

